# Head-to-head comparison of diagnostic accuracy of TB screening tests: Chest-X-ray, Xpert TB host response, and C-reactive protein

**DOI:** 10.1101/2024.06.20.24308402

**Authors:** Rebecca Crowder, Balamugesh Thangakunam, Alfred Andama, Devasahayam J Christopher, Victoria Dalay, Welile Dube-Nwamba, Sandra V. Kik, Dong Van Nguyen, Nguyen Viet Nhung, Patrick PJ Phillips, Morten Ruhwald, Grant Theron, William Worodria, Charles Yu, Payam Nahid, Adithya Cattamanchi, Ankur Gupta-Wright, Claudia M. Denkinger, R2D2 TB Network

## Abstract

**Background:** Accessible, accurate screening tests are necessary to advance tuberculosis (TB) case finding and early detection in high-burden countries. We compared the diagnostic accuracy of available TB triage tests.

**Methods:** We prospectively screened consecutive adults with ≥2 weeks of cough presenting to primary health centers in the Philippines, Vietnam, South Africa, Uganda, and India. All participants received the index tests: chest-X-ray (CXR), venous or capillary Cepheid Xpert TB Host Response (HR) testing, and point-of-care C-reactive protein (CRP) testing (Boditech iChroma II). CXR images were processed using computer-aided detection (CAD) algorithms. We assessed diagnostic accuracy against a microbiologic reference standard (sputum Xpert Ultra, culture). Optimal cut-points were chosen to achieve sensitivity ≥90% and maximize specificity. Two-test screening algorithms were considered, using two approaches: 1) sequential negative serial screening in which the second screening test is conducted only if the first is negative and positive is defined as positive on either test and 2) sequential positive serial screening, in which the second screening test is conducted only if the first is positive and positive is defined as positive on both tests.

**Results:** Between July 2021 and August 2022, 1,392 participants with presumptive TB had valid results on index tests and the reference standard, and 303 (22%) had confirmed TB. In head-to-head comparisons, CAD4TB v7 showed the highest specificity when using a cut-point that achieves 90% sensitivity (70.3% vs. 65.1% for Xpert HR, difference 95% CI 1.6 to 8.9; 49.7% for CRP, difference 95% CI 17.0 to 24.3). Among the possible two-test screening algorithms, three met WHO target product profile (TPP) minimum accuracy thresholds and had higher accuracy than any test alone. At 90% sensitivity, the specificity was 79.6% for Xpert HR-CAD4TB [sequential negative], 75.9% for CRP-CAD4TB [sequential negative], and 73.7% for Xpert HR-CAD4TB [sequential positive].

**Conclusions:** CAD4TB achieves TPP targets and outperforms Xpert HR and CRP. Combining screening tests further increased accuracy. Cost and feasibility of two-test screening algorithms should be explored.

**Registration:** NCT04923958

## INTRODUCTION

Approximately 7.5 million people were diagnosed with Tuberculosis (TB) in 2022 and 1.3 million people died from the disease [1]. Globally, 3.1 million TB cases were undiagnosed or unreported in 2022 [1]. This demonstrates that access to high-quality and affordable TB testing remains limited, making TB diagnosis the weakest link in the TB cascade of care. World Health Organization (WHO) target product profiles (TPPs) for TB diagnostics prioritize a simple, low-cost, non-sputum-based point-of-care (POC) triage or screening test to rule-out TB and guide confirmatory testing [2].

Promising TB triage tests include improvements on established technologies, such as chest X-ray (CXR); new application of existing tests, such as C-reactive protein (CRP); and novel assays, including the Xpert TB Host Response (Xpert HR) cartridge (Cepheid, Sunnyvale, USA). CXR implementation for TB screening has been limited in resource-constrained settings due to centralized CXR infrastructure [3] and scarcity of skilled personnel for interpretation [4]. The advent of digital CXR with computer-aided detection (CAD) analysis may reduce barriers to CXR screening by quickly enabling screening without the need for a skilled reader [5, 6]. However, performance varies by population screened, and limited data are available for children [7-10]. CRP is a non-specific marker for inflammation that has shown better accuracy than symptom screening for TB among people living with HIV (PLHIV) [11]. High CRP levels correlate with mycobacterial load and are associated with poor prognostic clinical features and a higher risk of death [12, 13]; however, CRP alone has not been shown to meet TPP specificity targets [11, 12, 14]. The recently developed Xpert HR assay detects expression levels of three host genes in whole blood. Results from early studies found that this assay approaches WHO minimum accuracy targets for a non-sputum-based triage test [15, 16].

Direct comparison of diagnostic accuracy, alongside comparisons of cost and operational characteristics, are needed to guide policy decisions around the implementation of novel TB screening tests and assess consistency with TPP targets. Testing algorithms that combine available tests may be needed to meet desired characteristics. CXR combined with CAD as a second screening test is considered a possible option by WHO. We aimed to conduct a head-to-head comparison of the diagnostic accuracy of three potential TB screening tests, including CAD, CRP, and Xpert HR in a large multi-country cohort of people with presumptive TB.

## METHODS

### Study design, population, and procedures

In this prospective study, we screened adults (≥18 years) presenting to primary health centers with presumed TB between July 2021 and August 2022 in the Philippines, Vietnam, Uganda, South Africa, and India. Specific enrollment locations have been reported previously [16] and are listed in the Appendix (**Appendix Table S1**). We enrolled consecutive outpatients if they had ≥2 weeks of new or worsening cough. We excluded people who had completed treatment for TB infection or disease within the past 12 months, had taken antibiotics with anti-mycobacterial activity within 2 weeks of study entry, or were unable or unwilling to return for follow-up or provide informed consent. All participants had HIV and diabetes screening (using HbA1c), and 2-3 spot sputum samples were collected for reference standard testing (sputum was induced if unable to expectorate spontaneously). Study participants received CXR, provided venous or capillary blood for Xpert HR testing, and provided capillary blood for CRP testing.

### Index testing

All participants underwent antero-posterior or postero-anterior CXR at baseline. If digital CXR was not available, analog CXR images were scanned to create jpeg images which were then converted into DICOM format using the img2dcm tool from the dcmtk toolkit (v3.6.6). Images that did not fulfill the DICOM features that were required for successful CAD software processing were subsequently modified using the dcmodify tool (v3.6.6) from the dcmtk toolkit before they were processed with the CAD software. Images were processed using five different CAD algorithms, but based on previous analysis [17], we chose the best performing CAD algorithm, CAD4TB v7, (CAD4TB), to be evaluated for comparisons in this analysis. CAD analysis was conducted by FIND, according to the developers’ instructions. CAD4TB produces a TB score, ranging from 0 to 100.

The Xpert® MTB-HR Research-Use-Only (RUO) prototype cartridge evaluates mRNA levels of four differentially expressed genes (GBP5, DUSP3, KLF2 and TBP). Testing involved inoculating the cartridge by transferring 100 µL of freshly collected whole or capillary blood in an EDTA tube into the cartridge chamber and analyzing using the GeneXpert instrument as per manufacturer instructions. We calculated the ‘TB score’ using the formula (Ct GBP5 + Ct DUSP3)/2 − Ct TBP as previously described [16].

CRP concentrations were measured at baseline from capillary blood using a United States Food and Drug Administration-approved standard sensitivity POC assay, measured using ichroma II (Boditech Med Inc., Chuncheon-si, South Korea) that provides quantitative results in three minutes (range: 2.5-300 mg/L). A fixed-volume micropipette (50 µL) was used to obtain and transfer capillary blood to a reagent tube, mixed, and two drops were applied on the lateral-flow device for testing.

No pre-specified cut-point was provided for any index test. All operators performing the index and reference tests were blinded to clinical details and other TB testing results.

### Reference standard testing

Each participant provided 2-3 sputum samples, and the microbiologic reference standard (MRS) was determined by results from sputum Xpert Ultra MTB/RIF (Cepheid, Sunnyvale, USA) and mycobacterial culture (MGIT, Becton Dickinson, USA; solid culture in 7h10 media was used when MGIT supplies were not available). A positive reference standard was defined as a positive sputum Xpert Ultra result (grade ‘very low’ or higher, or two ‘trace’ results) or a positive culture. Participants with a positive Xpert Ultra result did not undergo confirmatory culture. A negative reference standard was defined as no positive results on Xpert Ultra or culture and two negative cultures. Those not meeting the criteria for positive or negative were defined as indeterminate and excluded from the analysis.

In addition, we used a secondary, simplified sputum Xpert Ultra reference standard given that a molecular test is typically the only confirmatory TB test available in programmatic settings.

### Statistical Analysis

For each index test, we conducted receiver operating characteristic (ROC) analysis to compute the area under the curve (AUC) with 95% confidence interval (CI). Participants with indeterminate index test results were excluded from the analysis. For the primary analysis, optimal cut-points were chosen for each test that would achieve the TPP target sensitivity ≥90% and maximize specificity in the overall population. Two-test screening algorithms were also considered. We considered 1) a sequential negative serial screening approach [18], in which the second screening test is conducted only if the first is negative and a positive screen is defined as positive on either test and 2) a sequential positive serial screening approach [18], in which the second screening test is conducted only if the first is positive and a positive screen is defined as positive on both tests (**Appendix Figure S1**). We considered CRP and Xpert HR the more likely initial test, followed by the CAD, given the infrastructure requirements necessary for CXR and CAD. For each potential combination, we considered 1,000 possible combinations of cut-points (100 possible cut-points for the first test and 100 possible cut-points for the second test). We then identified all pairs of cut-points that would achieve sensitivity ≥90% and specificity ≥70%, then likewise selected the pair that achieved the highest possible specificity within these constraints. With all binary tests set to have a sensitivity of 90%, our primary analysis compared the specificity of each index alone and the possible combined two-test screening algorithms among the entire study population. The absolute difference in specificity for each pairwise comparison was calculated using McNemar’s test for paired proportions. Results are reported following STARD guidance [19]. Secondary analyses compared the sensitivity and specificity of the index tests alone and in combination among subgroups of interest (by country, HIV status, diabetes status). A sample size of 1,500 (300 per country) was pre-specified to achieve reasonable confidence interval widths between 5.2-9.4% depending on observed test sensitivity (80-95%) and 4.6%-5.6% depending on observed test specificity (60-80%) when assuming a TB prevalence of 20%. Stata version 18 was used for all analyses.

### Ethics

This study was registered with ClinicalTrials.gov (NCT04923958). Study procedures were approved by the institutional review board (IRB) of the University of California San Francisco, the University of Heidelberg, and by local IRBs at each enrollment site (**Appendix Table S1**).

### Funding

This work was supported by the United States National Institute of Allergy and Infectious Diseases [U01AI152087]. POC CRP test kits and ichroma II machines were donated by Boditech Med Inc, and Xpert HR test kits were donated by Cepheid. CAD was performed by FIND at no cost to the study. The installation and use of the CAD4TB software evaluated in this manuscript was provided free of charge by Delft Imaging to FIND. Study funders and product developers had no role in study design, data collection, data analysis, data interpretation, or writing.

## RESULTS

### Participants

Between July 2021 and August 2022, 2,046 adults were screened across clinical sites in five countries (the Philippines, Vietnam, Uganda, South Africa, and India). 1,666 (81%) met the criteria for inclusion in the study, of which 1,561 (94%) received all three tests. 31 (2%) CAD4TB readings, 27 (2%) Xpert HR results, and no CRP results were indeterminate or invalid. 1,392 (89%) participants had all three index tests and the microbiologic reference standard performed with valid results (**Figure 1**). Of the 1,392 assessable participants, 46% were female, the median age was 43, 14% were living with HIV (median CD4 count 378), and 14% were living with diabetes (median HbA1c 7.0) (**Table 1**). In total, 303 (22%) had microbiologically confirmed TB. However, characteristics varied by country (**Table 1**), with TB prevalence ranging from 9% in India and the Philippines to 38% in Vietnam.

**Table 1.** Demographic and clinical characteristics by country.

|  | Total<br>(N=1,392) | Philippines<br>(N=326) | Vietnam<br>(N=276) | Uganda<br>(N=267) | South Africa<br>(N=262) | India<br>(N=261) |
| --- | --- | --- | --- | --- | --- | --- |
| Age, mean ± SD (years) | 43 ± 16 | 40 ± 15 | 51 ± 16 | 34 ± 12 | 40 ± 12 | 48 ± 16 |
| Female | 633 (45%) | 181 (56%) | 108 (39%) | 112 (42%) | 125 (48%) | 107 (41%) |
| Body mass index (BMI), mean ± SD | 23 ± 5 | 24 ± 5 | 20 ± 3 | 22 ± 4 | 24 ± 7 | 23 ± 6 |
| Person living with HIV | 197 (14%) | 3 (1%) | 2 (1%) | 80 (30%) | 102 (40%) | 10 (4%) |
| CD4 count, median (IQR) | 378 (210-651) | 527 (184-812) | 547 (497-597) | 360 (222-634) | 335 (204-660) | 587 (220-692) |
| Person living with diabetes | 196 (14%) | 42 (13%) | 65 (24%) | 25 (9%) | 17 (6%) | 47 (18%) |
| Prior TB diagnosis | 271 (19%) | 43 (13%) | 58 (21%) | 32 (12%) | 100 (38%) | 38 (15%) |
| **Symptoms** |  |  |  |  |  |  |
| Cough ≥ 2 weeks | 1392 (100%) | 326 (100%) | 276 (100%) | 267 (100%) | 262 (100%) | 261 (100%) |
| Fever | 489 (35%) | 50 (15%) | 98 (36%) | 184 (69%) | 93 (35%) | 64 (25%) |
| Hemoptysis | 170 (12%) | 23 (7%) | 61 (22%) | 44 (16%) | 19 (7%) | 23 (9%) |
| Night sweats | 474 (34%) | 56 (17%) | 89 (32%) | 167 (63%) | 136 (52%) | 26 (10%) |
| Weight loss | 639 (46%) | 91 (28%) | 79 (29%) | 187 (70%) | 168 (64%) | 114 (44%) |
| Loss of appetite | 515 (37%) | 75 (23%) | 73 (26%) | 157 (59%) | 114 (44%) | 96 (37%) |
| **Reference standards** |  |  |  |  |  |  |
| Sputum Xpert Ultra Reference Standard |  |  |  |  |  |  |
| TB Negative | 1114 (80%) | 304 (93%) | 183 (66%) | 176 (66%) | 213 (81%) | 238 (91%) |
| TB Positive | 274 (20%) | 22 (7%) | 93 (34%) | 90 (34%) | 47 (18%) | 22 (8%) |
| Indeterminate | 4 (0%) | 0 (0%) | 0 (0%) | 1 (0%) | 2 (1%) | 1 (0%) |
| Highest semiquantitative grade |  |  |  |  |  |  |
| Trace | 6 (2%) | 1 (5%) | 0 (0%) | 3 (3%) | 2 (4%) | 0 (0%) |
| Very low | 43 (16%) | 5 (23%) | 14 (15%) | 10 (11%) | 10 (20%) | 4 (18%) |
| Low | 85 (31%) | 8 (36%) | 37 (40%) | 19 (21%) | 12 (24%) | 9 (41%) |
| Medium | 68 (25%) | 6 (27%) | 21 (23%) | 23 (25%) | 12 (24%) | 6 (27%) |
| High | 75 (27%) | 2 (9%) | 21 (23%) | 36 (40%) | 13 (27%) | 3 (14%) |
| Microbiologic reference standard |  |  |  |  |  |  |
| TB Negative | 1089 (78%) | 297 (91%) | 171 (62%) | 174 (65%) | 209 (80%) | 238 (91%) |
| TB Positive | 303 (22%) | 29 (9%) | 105 (38%) | 93 (35%) | 53 (20%) | 23 (9%) |
| **Sample collection** |  |  |  |  |  |  |
| Blood collection method |  |  |  |  |  |  |
| Venous | 1143 (82%) | 326 (100%) | 276 (100%) | 267 (100%) | 13 (5%) | 261 (100%) |
| Capillary | 249 (18%) | 0 (0%) | 0 (0%) | 0 (0%) | 249 (95%) | 0 (0%) |
| CXR type |  |  |  |  |  |  |
| Digital | 1148 (82%) | 326 (100%) | 276 (100%) | 23 (9%) | 262 (100%) | 261 (100%) |
| Analog | 244 (18%) | 0 (0%) | 0 (0%) | 244 (91%) | 0 (0%) | 0 (0%) |

Missing HIV-status for 6 participants in South Africa.

**Figure 1.**
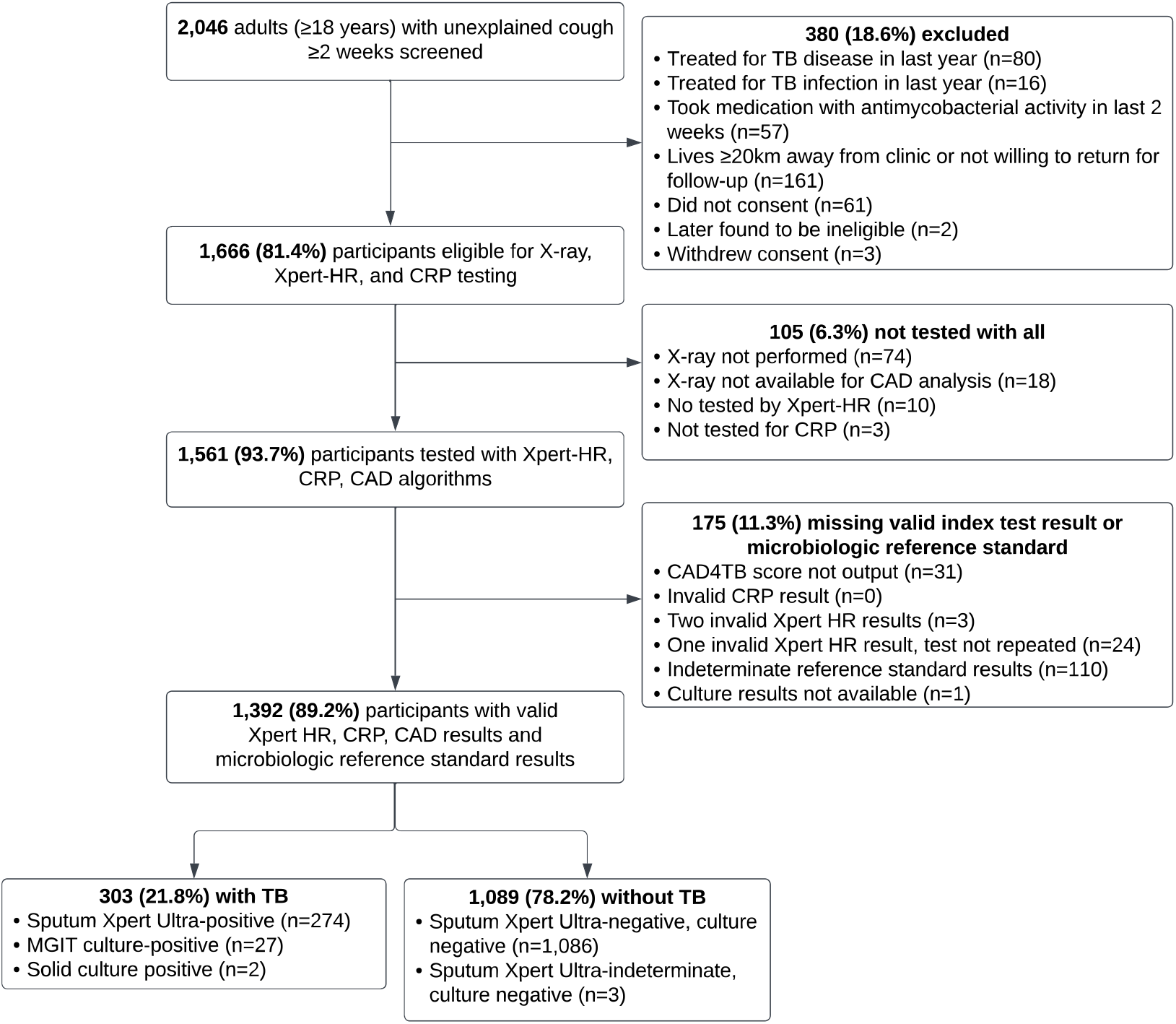
Study population. Note. Culture is not performed for participants with positive results on sputum Xpert Ultra.

Xpert HR was conducted using venous (82%, n=1,143) or capillary blood (18%, n=249). CXR images were collected digitally for 1,148 (83%), and analog images were scanned for 244 (18%). The distribution of each novel test quantitative result by microbiologic reference standard category is presented in the Appendix (**Appendix Table S2**). There were no adverse events from any testing.

The AUC for CAD4TB was 0.90 (95% CI 0.87, 0.92). Xpert HR had a similar AUC: 0.89 (95% CI 0.86, 0.91), and CRP only reached 0.81 (95% CI 0.79, 0.84) (**Figure 2**). ROC analysis against the sputum Xpert reference standard is available in the Appendix (**Figure S2**).

**Figure 2.**
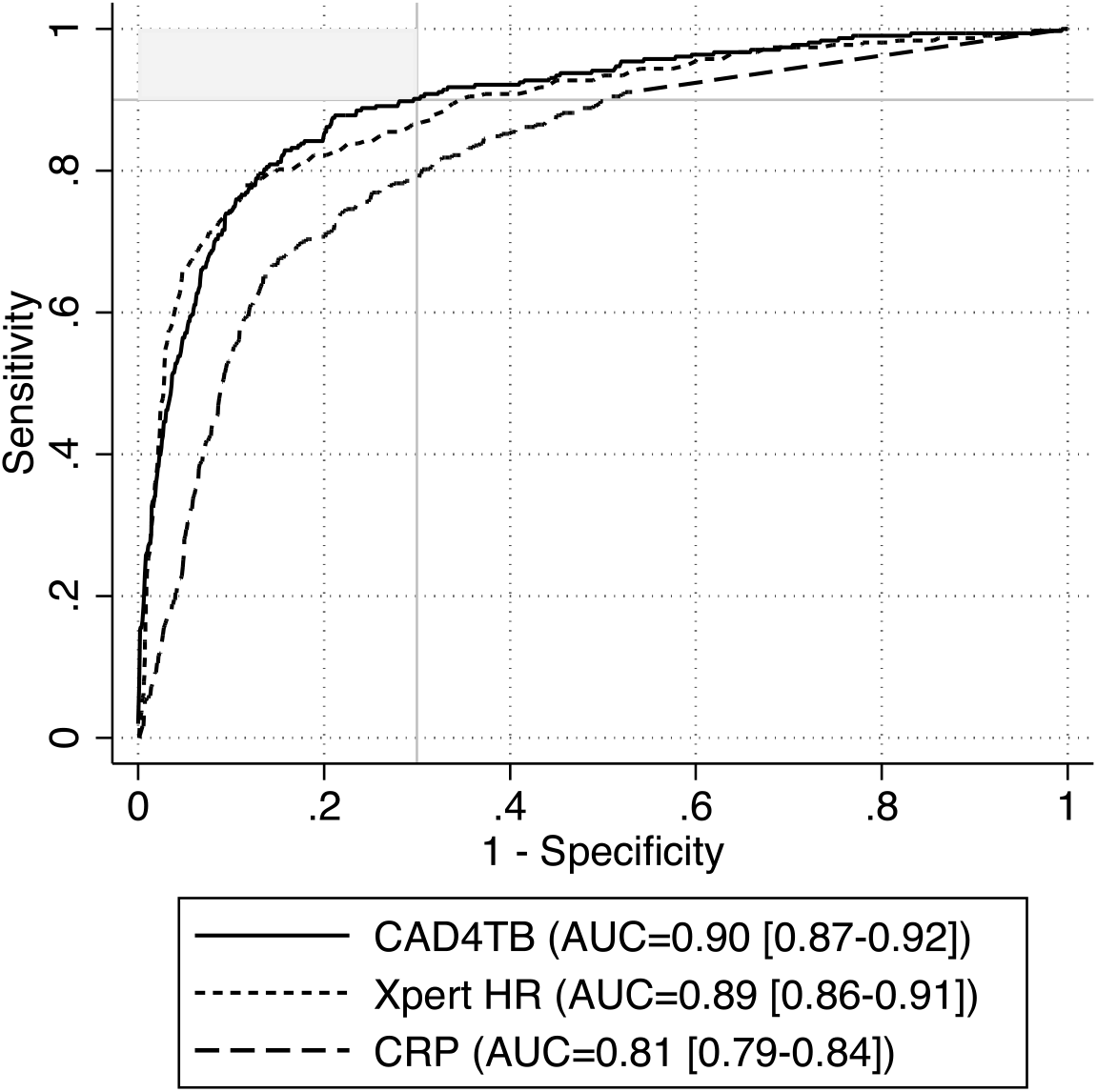
Receiver operating characteristic curve. ROC curves against the microbiologic reference standard with AUC and 95% CI displayed for CAD4TB, Xpert HR, and CRP. The upper-left area shaded in gray notes the region where tests meet TPP targets (≥90% sensitivity, ≥70% specificity). N=1,392 participants from the Philippines, Vietnam, Uganda, South Africa, and India with presumptive TB (n=303, 22% with microbiologically confirmed TB).

Against the microbiologic reference standard and at a sensitivity of 90%, CAD4TB (TB score ≥28.61) achieved the highest specificity 70.3% (95% CI 67.5, 73.0), and Xpert HR (TB score ≤-1.3) achieved a specificity of 65.1% (95% CI 62.2, 67.9). CRP with a cut-off of ≥2.81 mg/L achieved a specificity of 49.7% (95% CI 46.7, 52.7). At the WHO-recommended cut-point of ≥5mg/L, sensitivity of CRP was 82.8% (95% CI 78.1, 86.9) and specificity 64.8% (95% CI 61.9, 67.7).

Evaluating CAD4TB, Xpert HR, and CRP in comparison using these cut-points, CAD4TB performs best, with a specificity of 5.2% (95% CI 1.6, 8.9) greater than Xpert HR, and 20.6% (95% CI 17.0, 24.3) greater than CRP (**Table 2**). Accuracy against the sputum Xpert Ultra alone is presented in the Appendix (**Table S6**).

**Table 2.**
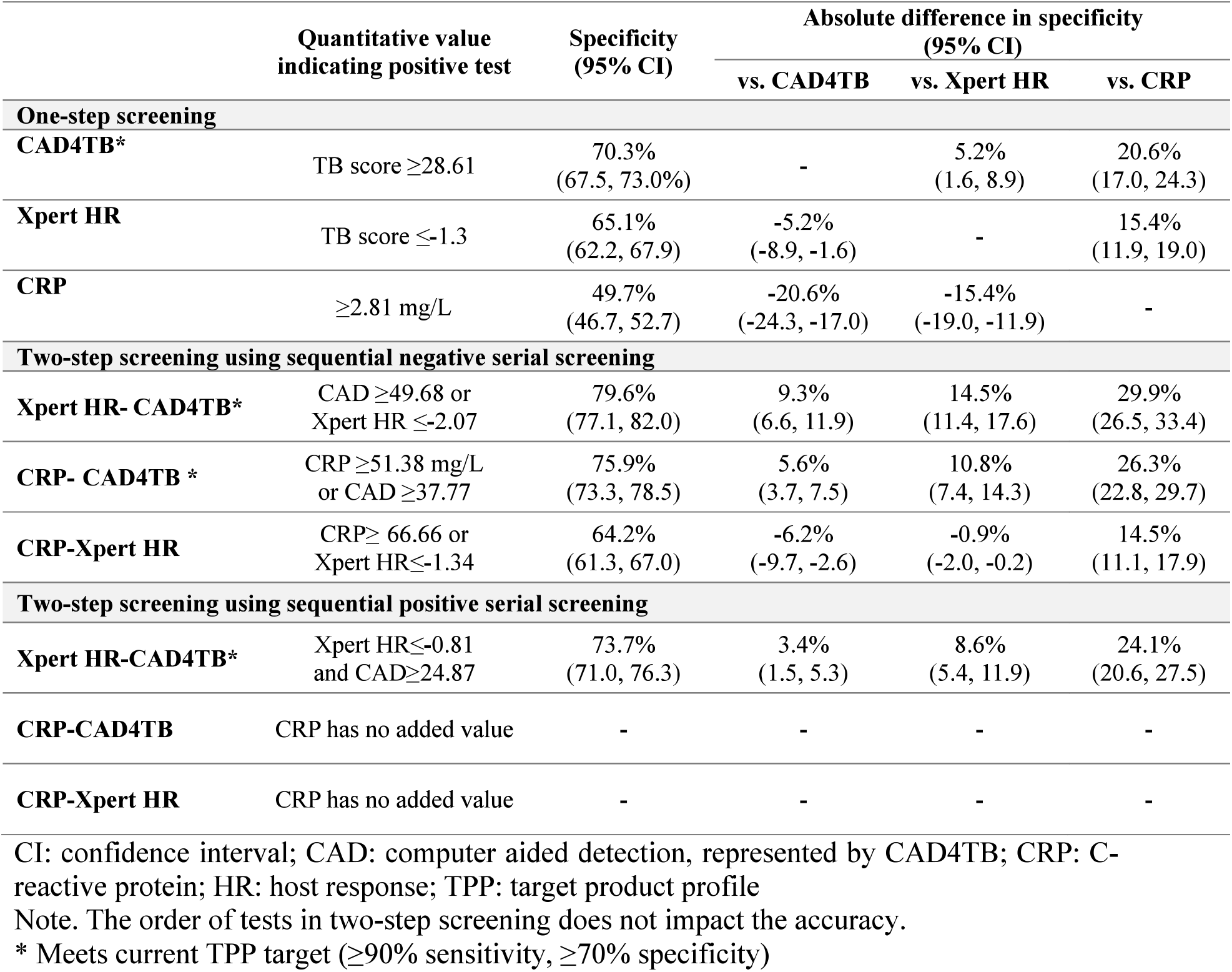
Head-to-head comparison of diagnostic accuracy at primary cut-points. Cut-points were chosen to achieve ≥90% sensitivity and maximize specificity against the microbiologic reference standard in the overall study population.

### Two-test screening algorithms

We found that 417 pairs of cut points for a combined Xpert HR-CAD4TB test achieved TPP targets when using a sequential negative serial screening approach, and 31 pairs of cut points achieved TPP targets using a sequential positive serial screening approach (**Figure 3**). The selection process for other two-test screening algorithms is presented in the Appendix (**Figure S3-S4**). The optimal combination of CAD4TB and Xpert HR (cut-point for CAD4TB ≥49.68 and Xpert HR ≤-2.07) achieved a specificity of 79.6% (95% CI 77.1, 82.0) with a sensitivity of 90%. This represents a 9.3% (95% CI 6.6, 11.9) improvement over CAD4TB alone, 14.5% improvement over Xpert HR alone (95% CI 11.4, 17.6), and 29.9% over CRP alone (95% CI 26.5, 33.4; **Table 2**). Other two-step screening algorithms had lower accuracy. Using sequential negative serial screening, both Xpert HR-CAD4TB and CRP-CAD4TB met current TPP targets. Using sequential positive serial screening, only Xpert HR-CAD4TB met the current TPP targets (**Table 2**). The agreement between tests and test algorithms is reported in the Appendix (**Table S7**).

**Figure 3.**
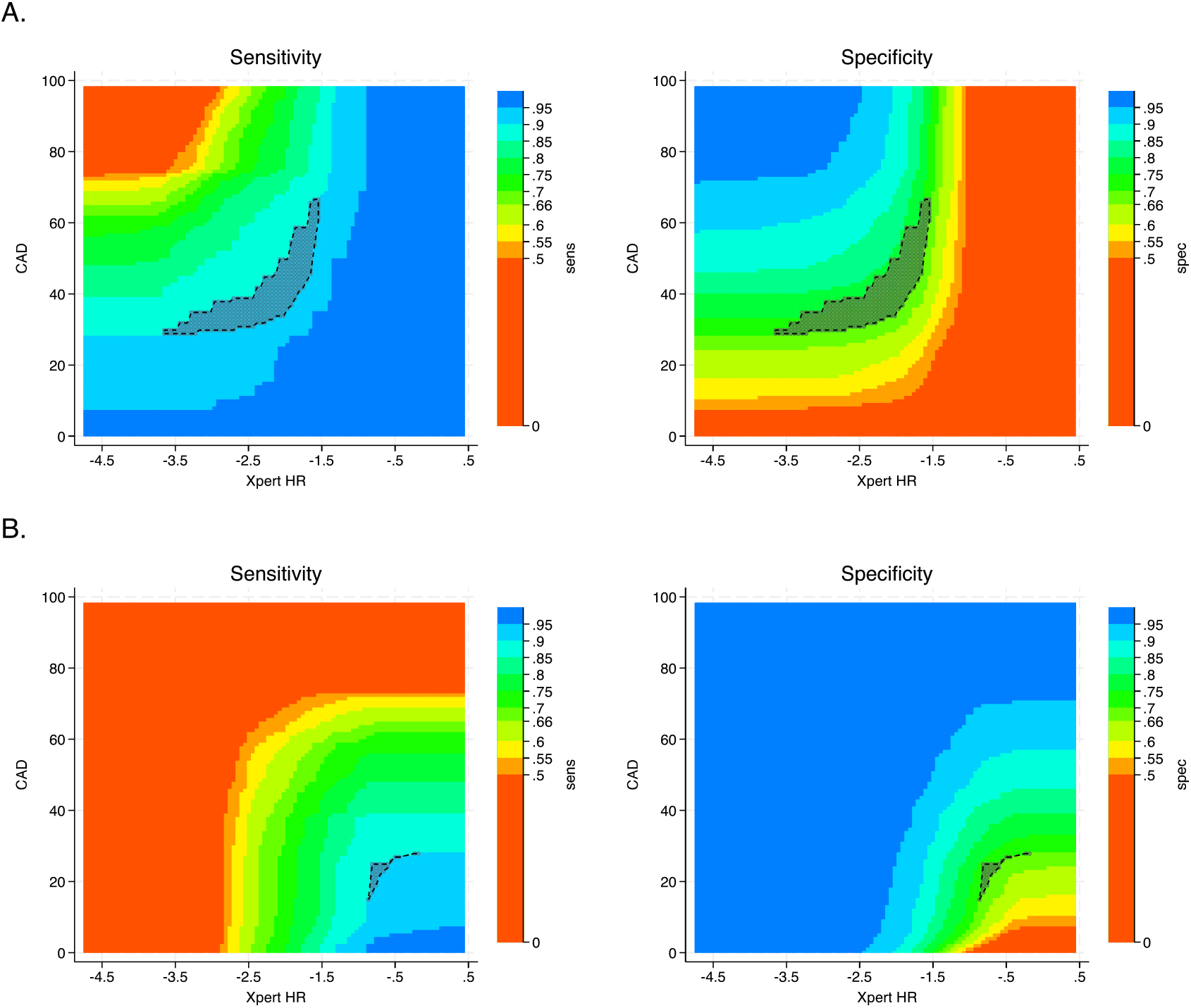
Selection of cut points for two-step screening algorithm combining Xpert HR and CAD4TB. Panel (A) shows the possible cut-points using sequential negative serial screening, in which the second screening test is conducted only if the first is negative and a positive screen is defined as positive on either test. Panel (B) shows the possible cut-points using sequential positive serial screening, in which the second screening test is conducted only if the first is positive and a positive screen is defined as positive on both tests. The x-axis shows all potential cut-points for Xpert HR (test positive defined as less than or equal to the cut point chosen), and the y-axis shows all potential cut points for CAD4TB (test positive defined as greater than or equal to the cut point chosen). Each point on the graph corresponds to a pair of cut points used to define a positive screening algorithm. The colors represent the range of sensitivities and specificities possible. The outlined region contains pairs with sensitivity≥90% and specificity ≥70% (n=417 in panel A, n=31 in panel B).

### Subgroup analyses

Using the same cut-points for the best-performing two-test screening algorithm (Xpert HR-CAD4TB using sequential negative serial screening), accuracy varied most notably by country, with sensitivity ranging from 62.1% (95% CI 42.3, 79.3) in the Philippines to 100% (95% CI 85.2, 100) in India. Sensitivity was similar among people with and without HIV (89.5% vs. 90.2%) and people with and without diabetes (sensitivity 91.7% vs. 89.7%); specificity was similar among people with and without diabetes (77.2% vs. 80.0%) and slightly higher among people without HIV (81.1% vs. 71.1% among PLHIV; **Appendix Figure S5**). Females had lower sensitivity (81.4% vs. 94.2% among males), but higher specificity (85.4% vs. 74.0%, **Appendix Figure S5**). Subgroup analyses of other combined tests are presented in the Appendix (**Figures S6-S8**).

Using subgroup-specific cut-points, all one- and two-step screening algorithms except CRP alone were able to achieve 90% sensitivity among participants in the Philippines (n/N=29/326 with MRS-positive TB) and among females (n/N=95/623 with MRS-positive TB). No screening algorithms met TPP targets among females, and only one screening algorithm, defined by Xpert HR≤-0.658 and CAD4TB≥6.727 met TPP targets among participants in the Philippines, with a specificity >70% (**Appendix Table S3**).

In a hypothetical cohort of 1,000 people with presumed TB, assuming a TB prevalence of 10%, the best-performing combined test would achieve a positive predictive value of 32.8% (95% CI 27.3, 38.8) and a negative predictive value of 98.6% (95% CI 97.5, 99.3) (**Appendix Table S4)**. Performing CAD4TB-Xpert HR using sequential negative serial screening (positive defined as CAD4TB ≥49.68 or Xpert HR ≤-2.07), followed by sputum testing if positive, would avert 726 sputum tests vs. 643 sputum tests averted if using CAD4TB alone (**Appendix Table S5**). Both screening strategies would miss 10 (10%) people with TB. However, to achieve this accuracy, performing CAD4TB-Xpert HR using sequential negative serial screening would require 73-77% of participants to receive a second screening test depending on order (**Appendix Table S5**).

## DISCUSSION

Comparing CAD4TB, Xpert HR, and CRP, we found that CAD4TBwas the most accurate TB triage test for pulmonary TB and the only test that met WHO TPP specificity targets (≥70% at ≥90% sensitivity) in a multi-country cohort of people with presumptive TB. Combining CAD4TB with Xpert HR or CRP using a sequential negative serial screening algorithm further improved accuracy, while only a combination with Xpert HR improved performance when using sequential positive serial screening. There was some variability in performance by subgroup; therefore, subgroup-specific cut-points may be warranted for best performance with implementation.

The diagnostic accuracy of CAD4TB in combination with other screening tests has not been well-studied, given the cost and infrastructure needs for CAD4TB alone are substantial. Nevertheless, per-screen costs of certain CAD solutions, when used at scale, have proven to be less costly compared with a radiologist [20] and more effective in terms of timeliness and completeness of results when implemented in a PLHIV population [21].

In a hypothetical cohort of 1,000 people with 10% TB prevalence, adding Xpert HR to CAD4TB using sequential negative serial screening averted 83 additional Xpert Ultra tests compared to using CAD4TB alone (274 vs. 357 requiring sputum Xpert testing); however, about three-quarters would require two screening tests prior to sputum testing (235 X-rays and CAD averted compared to CAD4TB alone). A sequential positive serial screening algorithm using the same tests would avert 288 X-rays compared to CAD4TB alone but require more confirmatory sputum testing (327 requiring sputum Xpert testing). Cost, however, is likely to be a major limitation of the implementation of a CAD4TB-Xpert HR screening approach, given that recent data showed the Xpert HR cartridge is unlikely to achieve cost-effectiveness if implemented alone [22]. While CRP-CAD would result in slightly decreased specificity using a sequential negative serial screening (3.7% less than Xpert HR-CAD4TB), it would avert 149 CAD4TB/CXR and 693 Xpert Ultra and thus warrants consideration from an economic standpoint. An analysis for Uganda of a CRP-CAD4TB algorithm suggested it could be costeffective [23].

Beyond cost, uptake of this intervention may be limited by access to the equipment needed (X-ray systems, GeneXpert instrument), especially at lower-level health facilities. Furthermore, the time it takes to complete screening using both screening tests may also reduce the effectiveness of this algorithm. A more cost-efficient two-stage screening algorithm would wait until the first test result (e.g., CRP or Xpert HR) comes back before running the second test (e.g., CAD4TB). This two-step approach is considered currently by the WHO in the revision of the TPP for TB screening. However, the added steps may further delay diagnosis or result in loss to follow-up, and this needs to be further studied with implementation [23].

We found that performance varied by country and sex could be mitigated by using subgroup-specific cut-points for test positivity. Tailoring guidelines for use will be important, as using universal cut-points may miss key subgroups. However, at the same time, different cut-offs might increase the complexity of validation for policy purposes and implementation.

Strengths of this study include its population: a large, well-characterized, diverse cohort of adults with presumptive TB across five countries. Prospective and concurrent evaluation of multiple TB triage tests allowed for a head-to-head comparison of these tests. This study is methodologically robust, with consistent implementation across sites and a thorough microbiological reference standard. However, several limitations should be considered in the interpretation of these findings: (1) The prevalence of TB differed across study sites, ranging from 9-38%, which may also suggest differing states of TB disease at the time of presentation across sites, which may explain at least some heterogeneity across sites; (2) limited generalizability beyond pulmonary TB; these results apply only to symptomatic pulmonary TB, and performance may differ in people unable to provide sputum and those with extrapulmonary TB.

In conclusion, current versions of CAD4TB achieve TPP targets for a TB triage test and outperform Xpert HR and CRP. Combining screening tests in algorithms further increases accuracy, but cost and implementation feasibility of these screening approaches will need to be evaluated.

## Supporting information

Additional contributors from the R2D2 TB Network

## Data Availability

The data that support the findings of this study are available from the corresponding author, CD, upon reasonable request.

## Author contributions

PN, AC, and CMD conceptualized the study. RC, PPJP, AC, AGW, and CMD designed the model framework. BT, VD, WD, AA, DVN, WW, CY, NVN, GT, and DJC collected the data. RC curated the data. RC, PPJP, and AGW conducted formal analysis. RC and PPJP visualized the data. RC, BT, WD, AA, AGW, and CMD drafted the manuscript. All authors edited and approved the manuscript.

## Notes

### Competing Interest Statement

The authors have declared no competing interest.

### Author Declarations

This study was registered with ClinicalTrials.gov (NCT04923958). Study procedures were approved by the institutional review board (IRB) of the University of California San Francisco, the University of Heidelberg, and by local IRBs at each enrollment site.

