## Additional contributors from the R2D2 TB Network for "Head-to-head comparison of diagnostic accuracy of TB screening tests: Chest-X-ray, Xpert TB host response, and C-reactive protein"

| First name | Surname | Affiliation |
|---|---|---|
| **India** | | |
| Shanmugasundaram | Elango | Christian Medical College, Vellore, India |
| Jerusha | Emmanuel | Christian Medical College, Vellore, India |
| Vinita | Ernest | Christian Medical College, Vellore, India |
| Priyadarshini | Gajendran | Christian Medical College, Vellore, India |
| Flavita | John | Christian Medical College, Vellore, India |
| Bharath | Karthikeyan | Christian Medical College, Vellore, India |
| Divya | Mangal | Christian Medical College, Vellore, India |
| Swetha | Sankar | Christian Medical College, Vellore, India |
| Rajasekar | Sekar | Christian Medical College, Vellore, India |
| Reena | Sekar | Christian Medical College, Vellore, India |
| Deepa | Shankar | Christian Medical College, Vellore, India |
| Mary | Shibiya | Christian Medical College, Vellore, India |
| Sai | Vijayasree | Christian Medical College, Vellore, India |
| **Philippines** | | |
| Jared | Almonte | De La Salle Medical and Health Sciences Institute, Cavite |
| Kevin Joshua | Alonzo | National TB Reference Laboratory, Research Institute for Tropical Medicine, Department of Health |
| Mary Faith | Angcaya | De La Salle Medical and Health Sciences Institute, Cavite |
| Joseph Edwin L. | Bascuña | National TB Reference Laboratory, Research Institute for Tropical Medicine, Department of Health |
| Ramon P. | Basilio | National TB Reference Laboratory, Research Institute for Tropical Medicine, Department of Health |
| Asella Ruvijean | Cariaga | De La Salle Medical and Health Sciences Institute, Cavite, Philippines |
| Gabriella | Castillon | De La Salle Medical and Health Sciences Institute, Cavite |
| Karlo | Dayawon | De La Salle Medical and Health Sciences Institute, Cavite |
| Raul | Destura | National Institutes of Health, University of the Philippines Manila |
| Jezreel | Esguerra | De La Salle Medical and Health Sciences Institute, Cavite |
| Eleonor | Garcia | De La Salle Medical and Health Sciences Institute, Cavite |
| Darecil | Gelina | De La Salle Medical and Health Sciences Institute, Cavite |
| Joseph Aldwin | Goleña | De La Salle Medical and Health Sciences Institute, Cavite |
| Maria Marissa | Golla | De La Salle Medical and Health Sciences Institute, Cavite |
| Emmanuelle | Gutierrez | De La Salle Medical and Health Sciences Institute, Cavite |
| Gidalthi Jonathan | Ilagan | De La Salle Medical and Health Sciences Institute, Cavite |
| Dodge R. | Lim | National TB Reference Laboratory, Research Institute for Tropical Medicine, Department of Health |
| Jaiem | Maranan | De La Salle Medical and Health Sciences Institute, Cavite |
| Danaida | Marcelo | De La Salle Medical and Health Sciences Institute, Cavite |
| Leonedy | Masangcay | De La Salle Medical and Health Sciences Institute, Cavite |
| Jenkin | Mendoza | National TB Reference Laboratory, Research Institute for Tropical Medicine, Department of Health |
| Angelita | Pabruada | De La Salle Medical and Health Sciences Institute, Cavite |
| Laarean | Perlas | De La Salle Medical and Health Sciences Institute, Cavite |
| Annalyn | Reyes | De La Salle Medical and Health Sciences Institute, Cavite |
| Roeus Vincent Arjay G. | Reyes | National TB Reference Laboratory, Research Institute for Tropical Medicine, Department of Health |

| First name | Surname | Affiliation |
|---|---|---|
| Lorenzo | Reyes | National TB Reference Laboratory, Research Institute for Tropical Medicine, Department of Health |
| Maria Guileane | Sanchez-Pogosa | National TB Reference Laboratory, Research Institute for Tropical Medicine, Department of Health |
| Maricef | Tonquin | De La Salle Medical and Health Sciences Institute, Cavite |
| **South Africa** | | |
| Shima | Abdulgadar | Stellenbosch University, Cape Town, South Africa |
| Cammy | Botha | Stellenbosch University, Cape Town, South Africa |
| Brigitta | Derendinger | Stellenbosch University, Cape Town, South Africa |
| Jane | Fortuin | Stellenbosch University, Cape Town, South Africa |
| Siphosethu | Gonya | Stellenbosch University, Cape Town, South Africa |
| Chumani | Hatile | Stellenbosch University, Cape Town, South Africa |
| Megan | Hendrikse | Stellenbosch University, Cape Town, South Africa |
| Charlotte | Lawn | Stellenbosch University, Cape Town, South Africa |
| Disha | Mathoorah | Stellenbosch University, Cape Town, South Africa |
| Desiree Lem | Mbu | Stellenbosch University, Cape Town, South Africa |
| Zintle | Ntetha | Stellenbosch University, Cape Town, South Africa |
| Anna | Okunola | Stellenbosch University, Cape Town, South Africa |
| Zaida | Palmer | Stellenbosch University, Cape Town, South Africa |
| Fikiswa | Seti | Stellenbosch University, Cape Town, South Africa |
| Charmaine | Van Der Walt | Stellenbosch University, Cape Town, South Africa |
| Lusanda | Yekani | Stellenbosch University, Cape Town, South Africa |
| **Uganda** | | |
| chriLucy | Asege | Walimu, Kampala, Uganda |
| Alice | Bukirwa | Walimu, Kampala, Uganda |
| David | Katumba | Walimu, Kampala, Uganda |
| Esther | Kisakye | Walimu, Kampala, Uganda |
| Wilson | Mangeni | Walimu, Kampala, Uganda |
| Job | Mukwatamundu | Walimu, Kampala, Uganda |
| Sandra | Mwebe | Walimu, Kampala, Uganda |
| Annet | Nakaweesa | Walimu, Kampala, Uganda |
| Martha | Nakaye | Walimu, Kampala, Uganda |
| Talemwa | Nalugwa | Walimu, Kampala, Uganda |
| Irene | Nassuna | Walimu, Kampala, Uganda |
| Irene | Nekesa | Walimu, Kampala, Uganda |
| Justine | Nyawere | Walimu, Kampala, Uganda |
| John Baptist | Ssonko | Walimu, Kampala, Uganda |
| **Vietnam** | | |
| Hai | Dang | Vietnam National Tuberculosis Program-University of California San Francisco Research Collaboration Unit; Center for Promotion of Advancement of Society, Hanoi, Vietnam |
| Luong | Dinh | Vietnam National Lung Hospital |
| Hang | Do | Hanoi Lung Hospital, Hanoi, Vietnam |
| Tam | Do | Hanoi Lung Hospital, Hanoi, Vietnam |
| Thuong | Do | Vietnam National Lung Hospital |
| Dung | Dao | Hanoi Lung Hospital, Hanoi, Vietnam |
| Ha | Doan | National TB reference Lab/ Vietnam National Lung Hospital, Hanoi, Vietnam |

| First name | Surname | Affiliation |
|---|---|---|
| Thien | Doan | Hanoi Lung Hospital, Hanoi, Vietnam |
| Huy | Ha | Vietnam National Tuberculosis Program-University of California San Francisco Research Collaboration Unit, Center for Promotion of Advancement of Society, Hanoi, Vietnam |
| Oanh | Lai | Hanoi Lung Hospital, Hanoi, Vietnam |
| Hien | Le | Vietnam National Tuberculosis Program-University of California San Francisco Research Collaboration Unit; Center for Promotion of Advancement of Society, Hanoi, Vietnam |
| Nguyet | Le | National TB reference Lab/ Vietnam National Lung Hospital, Hanoi, Vietnam |
| Anh | Nguyen | Hanoi Lung Hospital, Hanoi, Vietnam |
| Hanh | Nguyen | Vietnam National Tuberculosis Program-University of California San Francisco Research Collaboration Unit; Center for Promotion of Advancement of Society, Hanoi, Vietnam |
| Hoa | Nguyen | Vietnam National Lung Hospital |
| Hoang | Nguyen | Hanoi Lung Hospital, Hanoi, Vietnam |
| Thanh | Nguyen | Hanoi Lung Hospital, Hanoi, Vietnam |
| Yen | Nguyen | Hanoi Lung Hospital, Hanoi, Vietnam |
| Ha | Phan | Vietnam National Tuberculosis Program-University of California San Francisco Research Collaboration Unit, Center for Promotion of Advancement of Society, Hanoi, Vietnam |
| Nam | Pham | Vietnam National Tuberculosis Program-University of California San Francisco Research Collaboration Unit, Hanoi Lung Hospital, Hanoi, Vietnam |
| Thuong | Pham | Hanoi Lung Hospital, Hanoi, Vietnam |
| Trang | Trinh | Vietnam National Tuberculosis Program-University of California San Francisco Research Collaboration Unit, Center for Promotion of Advancement of Society, Hanoi, Vietnam |
| Phuong | Vu | Hanoi Lung Hospital, Hanoi, Vietnam |
| Trung | Vu | National TB reference Lab/ Vietnam National Lung Hospital, Hanoi, Vietnam |
| **USA** | | |
| Robert | Castro | University of California San Francisco, San Francisco, CA, USA |
| Adithya | Cattamanchi | University of California Irvine, Irvine, CA, USA |
| Catherine | Cook | University of California San Francisco, San Francisco, CA, USA |
| Sophie | Huddart | University of California San Francisco, San Francisco, CA, USA |
| Devan | Jaganath | University of California San Francisco, San Francisco, CA, USA |
| Midori | Kato-Maeda | University of California San Francisco, San Francisco, CA, USA |
| Tessa | Mochizuki | University of California San Francisco, San Francisco, CA, USA |
| Ruvandhi | Nathavitharana | Beth Israel Deaconess Medical Center, Harvard Medical School, Boston, MA, USA |
| Payam | Nahid | University of California San Francisco, San Francisco, CA, USA |
| Kevin | Nolan | University of California San Francisco, San Francisco, CA, USA |
| Kinari | Shah | University of California San Francisco, San Francisco, CA, USA |
| Swati | Sudarsan | University of California San Francisco, San Francisco, CA, USA |
| Christina | Yoon | University of California San Francisco, San Francisco, CA, USA |
| **Germany** | | |
| Maria del Mar | Castro Noriega | Heidelberg University Hospital, Heidelberg, Germany |

| First name | Surname | Affiliation |
|---|---|---|
| Theresa | Pfurtscheller | Heidelberg University Hospital, Heidelberg, Germany |
| Seda | Yerlikaya | Heidelberg University Hospital, Heidelberg, Germany |
| **Switzerland** | | |
| Matthew | Arentz | FIND, Geneva, Switzerland |
| Nathalie | Frey | FIND, Geneva, Switzerland |
| Sam | Linsen | FIND, Geneva, Switzerland |
