## Supplement for "Head-to-head comparison of diagnostic accuracy of TB screening tests: Chest-X-ray, Xpert TB host response, and C-reactive protein"

**APPENDIX**

**Table S1. R2D2 TB Network study enrolment sites and ethics committees**

| City, Country | Enrollment sites | Ethics Committee* |
|---|---|---|
| Vellore, India | CMC Pulmonary Outpatient Department, Primary care clinics in Vellore (Shalom/LCC, Chittor, CHAD) and Chitoor (CMC satellite campus) | Christian Medical College Institutional Review Board (13256) |
| Hanoi, Vietnam | Outpatient departments, Hanoi Lung Hospital | Ministry of Health Ethical Committee for National Biological Medical Research (94/CN-HĐĐĐ); National Lung Hospital Ethical Committee for Biological Medical Research (566/2020/NCKH); Hanoi Lung Hospital Science and Technology Initiative Committee (22/BVPHN) |
| Dasmariñas City, Philippines | Community-based screening in Dasmariñas City and nearby municipalities, outpatient clinics in Dasmariñas City | De La Salle Health Sciences Institute Independent Ethics Committee (2020-33-02-A) |
| Cape Town, South Africa | Scottsdene and Wallacedene primary care clinics; Brooklyn Chest Hospital; Khayelitsha District Health Center; Kraaifontein Community Health Clinic | Stellenbosch University Health Research Ethics Committee (M20/07/020) |
| Kampala, Uganda | Mulago Outpatient Department, Kisenyi Health Center, | Makerere University, College of Health Sciences, School of Medicine, Research Ethics Committee (2020-182) |

* The study was additionally approved by the University of California San Francisco Institutional Review Board (20-32670), and the University of Heidelberg Ethics Committee of the Medical Faculty (S-539/2020)

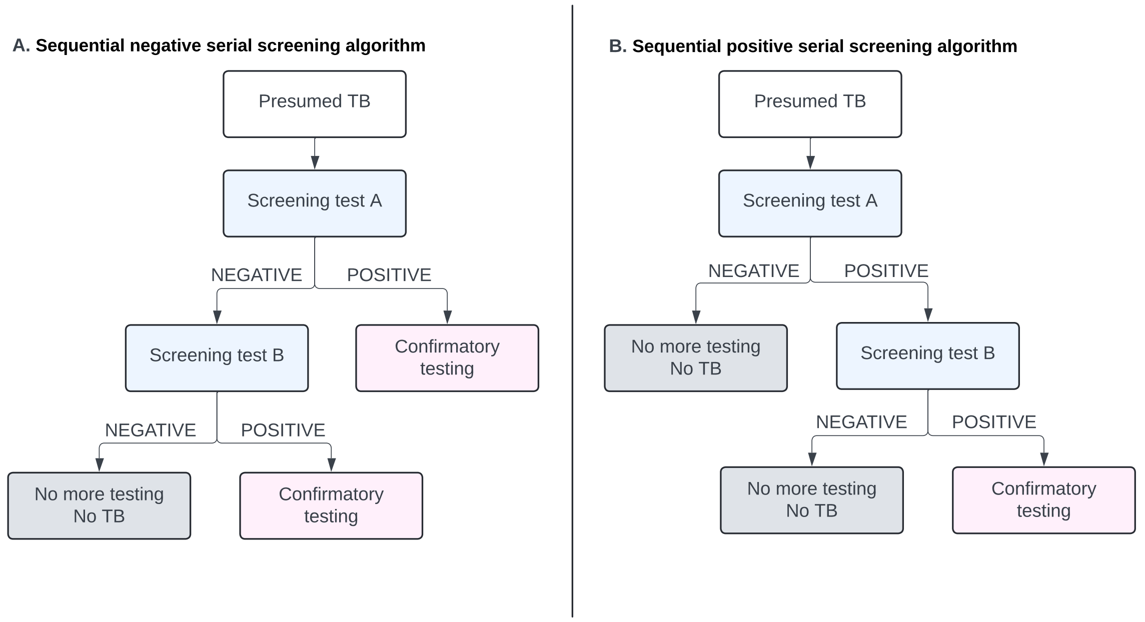

**Figure S1. Sequential testing algorithms.**
For each potential combination of screening tests, we considered a sequential negative serial screening approach (panel A) and a sequential positive serial screening approach (panel B).

**Table S2. Summary of index test results by reference standard classification**

| **Microbiologic Reference Standard** | | |
|---|---|---|
| **Index test, mean (SD)** | **Positive (n=303)** | **Negative (n=1,089)** |
| CRP | 61.0 (65.3) | 15.6 (37.4) |
| Xpert HR | -2.67 (0.94) | -1.15 (0.70) |
| CAD4TB | 68.2 (26.0) | 20.1 (23.2) |

| **Sputum Xpert Reference Standard** | | | |
|---|---|---|---|
| **Index test, mean (SD)** | **Positive (n=274)** | **Negative (n=1,114)** | **Indeterminate (n=4)** |
| CRP | 65.3 (65.9) | 15.6 (37.5) | 43.6 (36.2) |
| Xpert HR | -2.79 (0.88) | -1.16 (0.70) | -2.19 (1.13) |
| CAD4TB | 69.6 (25.1) | 20.9 (24.0) | 46.1 (39.2) |

SD: standard deviation

**Table S3. Results using subgroup-specific cut-points**

| | Using Philippines-specific cut-points (N=326 people in the Philippines, 29 [9%] with TB) | | Using female-specific cut-points (N=623 females, 95 [15.3%] with TB) | |
|---|---|---|---|---|
| | Quantitative value indicating positive test | Specificity (95% CI) | Quantitative value indicating positive test | Specificity (95% CI) |
| **One-step screening** | | | | |
| **CAD4TB** | TB score ≥7.1697 | 61.6% (55.8, 67.2) | TB score≥6.005 | 53.7% (49.4, 58.0) |
| **Xpert HR** | TB score ≤-0.8499 | 45.5% (39.7, 51.3) | TB score≤1.15 | 54.1% (49.8, 58.4) |
| **CRP** | Did not achieve 90% sensitivity | | Did not achieve 90% sensitivity | |
| **Two-step screening using either test positive approach** | | | | |
| **Xpert HR-CAD4TB** | Xpert HR ≤-2.69 or CAD4TB score ≥6.73 | 60.3% (54.5, 65.9) | Xpert HR ≤-1.318 or CAD4TB score ≥62.393 | 63.8% (59.6, 67.9) |
| **CRP-CAD4TB** | CRP≥10.79 or CAD4TB score≥10.49 | 65.0% (59.3, 70.4) | CRP≥36.105 or CAD4TB score≥10.988 | 61.9% (57.7, 66.1) |
| **CRP-Xpert HR** | CRP≥23.22 or Xpert HR≤-0.81 | 45.5% (39.7, 51.3) | CRP≥23.224 or Xpert HR≤-1.116 | 53.7% (49.4, 58.0) |
| **Two-step screening using both test positive approach** | | | | |
| **Xpert HR-CAD4TB*** | Xpert HR≤-0.658 and CAD4TB≥6.727 | 71.4% (65.9, 76.5) | Xpert HR≤-1.015 and CAD4TB≥3.079 | 63.2% (59.0, 67.3) |
| **CRP-CAD4TB** | CRP has no added value | - | - | - |
| **CRP-Xpert HR** | CRP has no added value | - | - | - |

CI: confidence interval; CAD: computer aided detection; CRP: C-reactive protein; HR: host response; TPP: target product profile

* Meets current TPP target in the Philippines (≥90% sensitivity, ≥70% specificity)

^ Meets current TPP target among females (≥90% sensitivity, ≥70% specificity)

**Table S4. Positive predictive value and negative predictive value in a hypothetical cohort of 1,000 with 10% TB prevalence**

| | Positive predictive value (95% CI) n=100 people with TB | Negative predictive value (95% CI) (n=900 people without TB) |
|---|---|---|
| **One-step screening** | | |
| CAD4TB | 25.2% (20.8, 30.0) | 98.4% (97.2, 99.3) |
| Xpert HR | 22.3% (18.3, 26.7) | 98.3% (96.9, 99.2) |
| CRP | 16.6% (13.5, 20.0) | 97.8% (96.0, 98.9) |
| **Two-step screening using either test positive approach** | | |
| Xpert HR-CAD4TB* | 32.8% (27.3, 38.8) | 98.6% (97.5, 99.3) |
| CRP-CAD4TB* | 29.3% (24.3, 34.8) | 98.6% (97.4, 99.3) |
| CRP-Xpert HR | 21.8% (17.9, 26.2) | 98.3% (96.9, 99.2) |
| **Two-step screening using both test positive approach** | | |
| Xpert HR-CAD4TB* | 27.5% (22.8, 32.7) | 98.5% (97.3, 99.3) |

CI: confidence interval

**Table S5. The potential number of sputum tests averted by using a triage test in a hypothetical cohort of 1000 with 10% TB prevalence**

| | **Number that have first test performed** (total population) | **Number that have second test performed** | **Number of people with TB missed** (n=100 with TB) | **Number of people correctly classified by triage test** | **Number of people triage test positive, requiring sputum testing** (N=1000 with presumptive TB) | **Number of sputum tests averted** (N=1000 with presumptive TB) |
|---|---|---|---|---|---|---|
| **One-step screening** | | | | | | |
| CAD4TB | 1,000 | 0 | 10 (10%) | 723 (72.3%) | 357 (35.7%) | 643 |
| Xpert HR | 1,000 | 0 | 10 (10%) | 676 (67.6%) | 404 (40.4%) | 596 |
| CRP | 1,000 | 0 | 10 (10%) | 537 (53.7%) | 543 (54.3%) | 457 |
| **Two-step screening using either test positive approach** | | | | | | |
| Xpert HR-CAD4TB*<br>CAD4TB-Xpert HR* | 1,000 | 765<br>728 | 10 (10%) | 806 (80.6%) | 274 (27.4%) | 726 |
| CRP-CAD4TB*<br>CAD4TB-CRP* | 1,000 | 851<br>643 | 10 (10%) | 773 (77.3%) | 307 (30.7%) | 693 |
| CRP-Xpert HR<br>Xpert HR-CRP | 1,000 | 884<br>541 | 10 (10%) | 668 (66.8%) | 412 (41.2%) | 588 |
| **Two-step screening using both test positive approach** | | | | | | |
| Xpert HR-CAD4TB*<br>CAD4TB-Xpert HR* | 1,000 | 712<br>463 | 10 (10%) | 753 (75.3%) | 327 (32.7%) | 673 |
| CAD4TB-CRP | 1,000 | 435 | 10 (10%) | 716 (71.6%) | 364 (36.4%) | 636 |
| Xpert HR-CRP | 1,000 | 476 | 10 (10%) | 670 (67%) | 410 (41%) | 590 |

Note. The order of tests in two-step screening does not impact the accuracy.

* Meets current TPP target (≥90% sensitivity, ≥70% specificity)

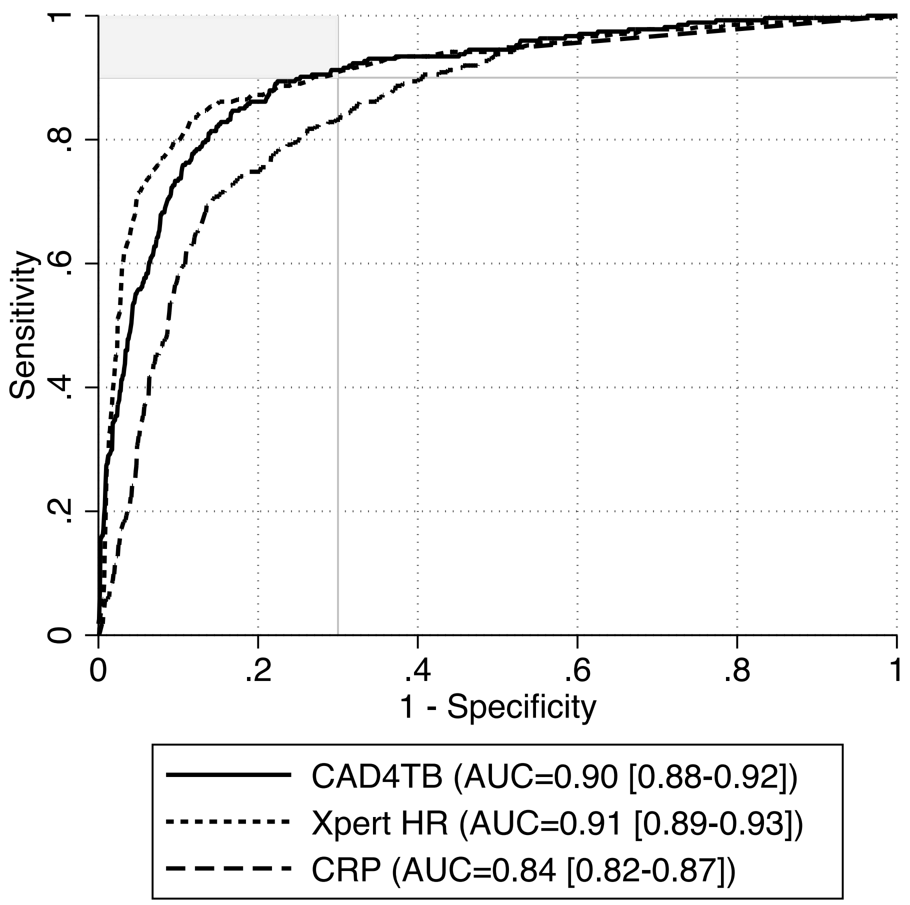

**Figure S2. Sputum Xpert reference standard receiver operating characteristic curve.** ROC curves with AUC and 95% CI displayed for CAD4TB, Xpert HR, and CRP. The upper-left area shaded in gray notes the region where tests meet TPP targets (≥90% sensitivity, ≥70% specificity). N=1,388 participants from the Philippines, Vietnam, Uganda, South Africa, and India with presumptive TB (n=274, 20% with sputum Xpert-positive TB). 4 participants included in the primary analysis had an indeterminate sputum Xpert Ultra result and were excluded from this analysis.

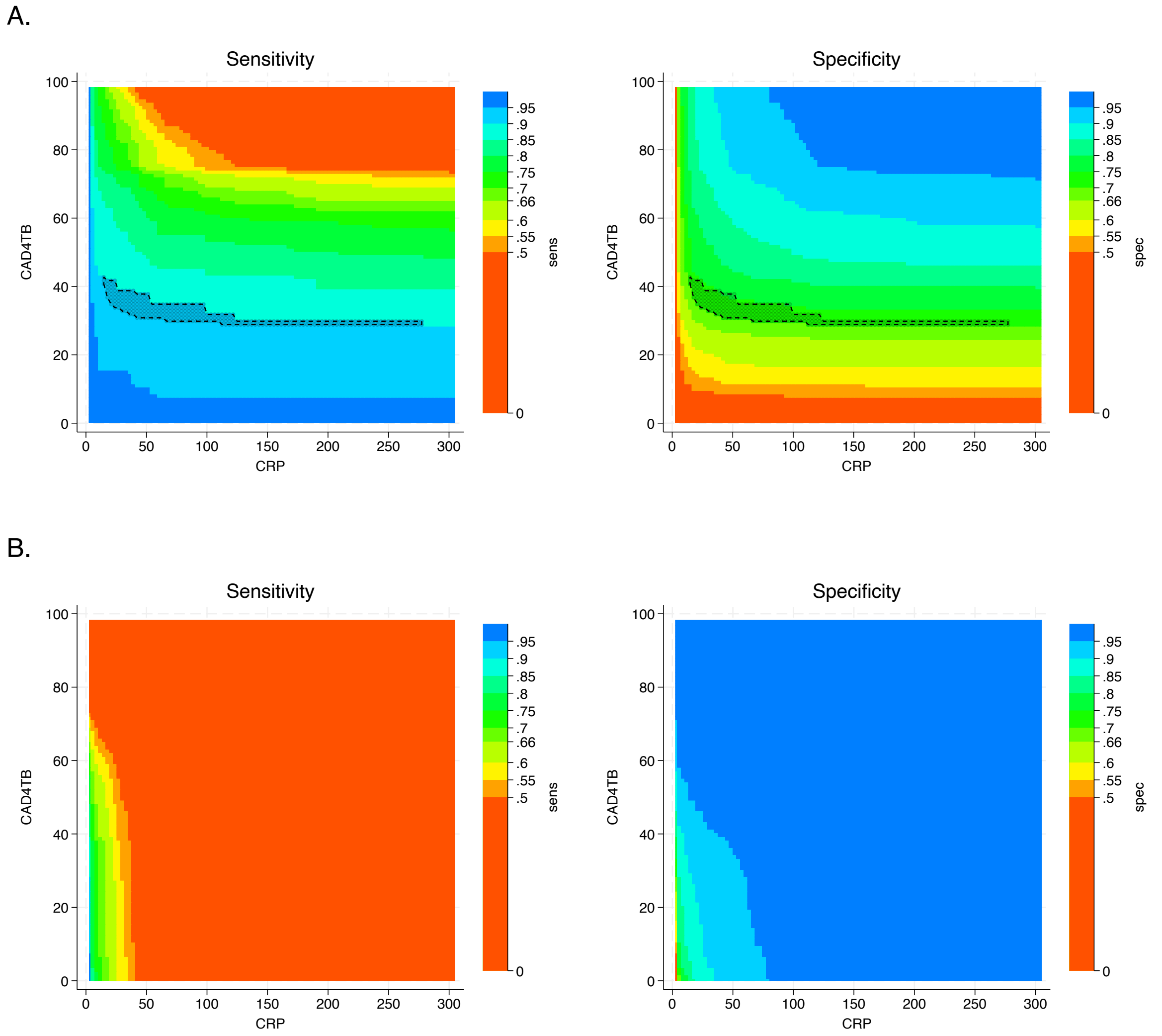

**Figure S3. Selection of cut points for two-step screening algorithm combining CRP and CAD4TB.** Panel (A) shows the possible cut-points using the sequential negative serial screening approach, in which the second screening test is conducted only if the first is negative and a positive screen is defined as positive on either test. Panel (B) shows the possible cut-points using the sequential positive serial screening approach, in which the second screening test is conducted only if the first is positive and a positive screen is defined as positive on both tests. The x-axis shows all potential cut-points for CAD4TB (test positive defined as greater than or equal to the cut point chosen), and the y-axis shows all potential cut points for CRP (test positive defined as greater than or equal to the cut point chosen). Each point on the graph corresponds to a pair of cut points used to define a positive screening algorithm. The colors represent the range of sensitivities and specificities possible. The outlined region contains pairs with sensitivity≥90% and specificity ≥70% (n=310 in panel A, n=0 in panel B).

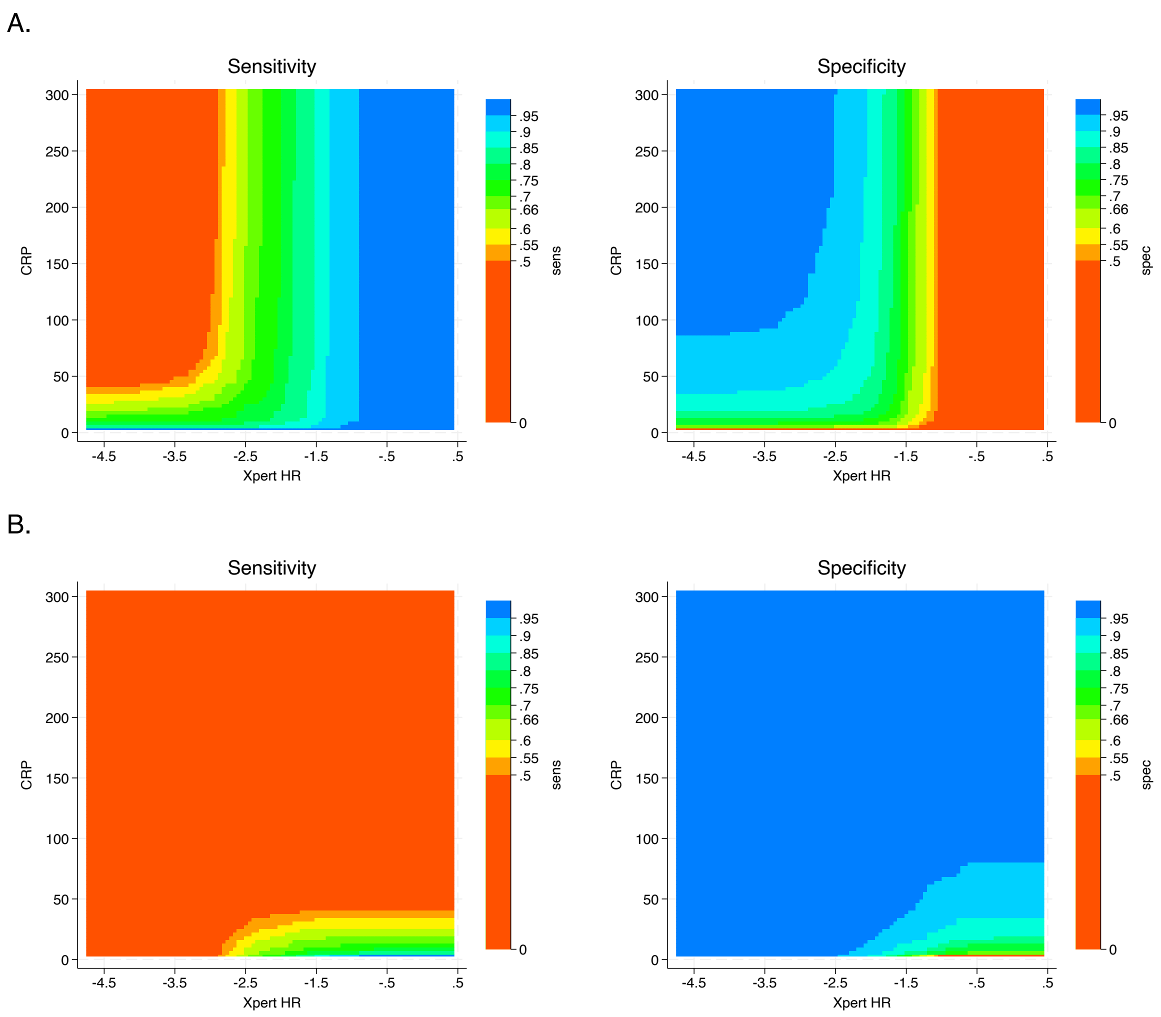

**Figure S4. Selection of cut points for two-step screening algorithm combining CRP and Xpert HR.** Panel (A) shows the possible cut-points using the sequential negative serial screening approach, in which the second screening test is conducted only if the first is negative and a positive screen is defined as positive on either test. Panel (B) shows the possible cut-points using the sequential positive serial screening approach, in which the second screening test is conducted only if the first is positive and a positive screen is defined as positive on both tests. The x-axis shows all potential cut-points for Xpert HR (test positive defined as less than or equal to the cut point chosen), and the y-axis shows all potential cut points for CRP (test positive defined as greater than or equal to the cut point chosen). Each point on the graph corresponds to a pair of cut points used to define a positive screening algorithm. The colors represent the range of sensitivities and specificities possible. No pair of cut points using either approach achieved sensitivity≥90% and specificity ≥70%.

**Table S6. Head-to-head comparison of diagnostic accuracy against sputum Xpert reference standard.** Cut points were chosen to achieve ≥90% sensitivity and maximize specificity against the sputum Xpert reference standard.

| N=1,388 | Quantitative value indicating positive test | Specificity (95% CI) | Absolute difference in specificity (95% CI) | | |
|---|---|---|---|---|---|
| | | | vs. CAD4TB | vs. Xpert HR | vs. CRP |
| **One-step screening** | | | | | |
| **CAD4TB*** | TB score ≥34.08 | 74.7% (72.0, 77.2) | - | 1.7 (-1.7, 5.1) | 15.4 (11.9, 18.8) |
| **Xpert HR*** | TB score ≤-1.499 | 73.0% (70.3, 75.6) | -1.7 (-5.1, 1.7) | - | 13.6% (10.3, 17.0) |
| **CRP** | ≥4.07 mg/L | 59.3% (56.4, 62.2) | -15.4 (-18.8, -11.9) | -13.6% (-17.0, -10.3) | - |
| **Two-step screening using either test positive appraoch** | | | | | |
| **Xpert HR-CAD4TB*** | CAD4TB ≥62.59 or Xpert HR ≤-2.28 | 85.1% (82.9, 87.1) | 10.4 (7.9, 12.9) | 12.1 (9.4, 14.9) | 25.8 (22.6, 29.0) |
| **CRP-CAD4TB*** | CRP ≥45.71 mg/L or CAD4TB ≥39.16 | 78.0% (75.5, 80.4) | 3.3 (1.3, 5.3) | 5.0 (1.8, 8.3) | 18.7 (15.5, 21.8) |
| **CRP-Xpert HR*** | CRP ≥265.23mg/L or Xpert HR ≤-1.495 | 72.9% (70.2, 75.5) | -1.8 (-5.2, 1.6) | 0.1 (-0.4, 0.2) | 13.6 (10.2, 16.9) |
| **Two-step screening using both test positive approach** | | | | | |
| **Xpert HR-CAD4TB*** | CAD4TB ≥31.82 and Xpert HR ≤-0.76 | 77.6% (75.1, 80.1) | 3.0 (1.6, 4.3) | 4.7 (1.5, 7.9) | 18.3 (15.0, 21.6) |
| **CRP-CAD4TB** | CRP has no added value | - | - | - | - |
| **CRP-Xpert HR** | CRP has no added value | - | - | - | - |

CI: confidence interval; CAD: computer aided detection, represented by CAD4TB; CRP: C-reactive protein; HR: host response; TPP: target product profile

N=1,392 people with presumptive TB, n=274 with sputum Xpert Ultra positive TB
Note. 4 participants included in the primary analysis had an indeterminate sputum Xpert Ultra result and were excluded from this analysis.
* Meets current TPP target (≥90% sensitivity, ≥70% specificity)

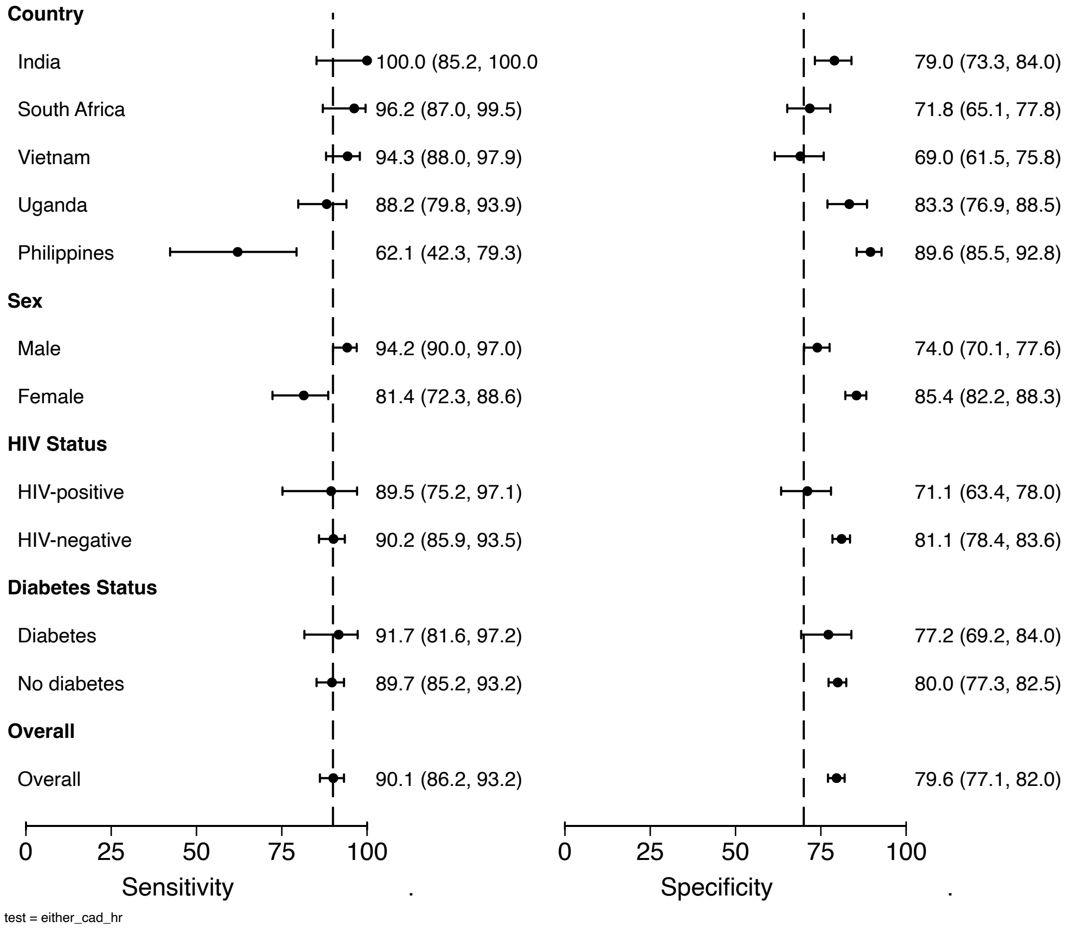

**Figure S5. Subgroup analysis of two-step screening algorithm Xpert HR-CAD4TB using a sequential negative serial screening algorithm.**

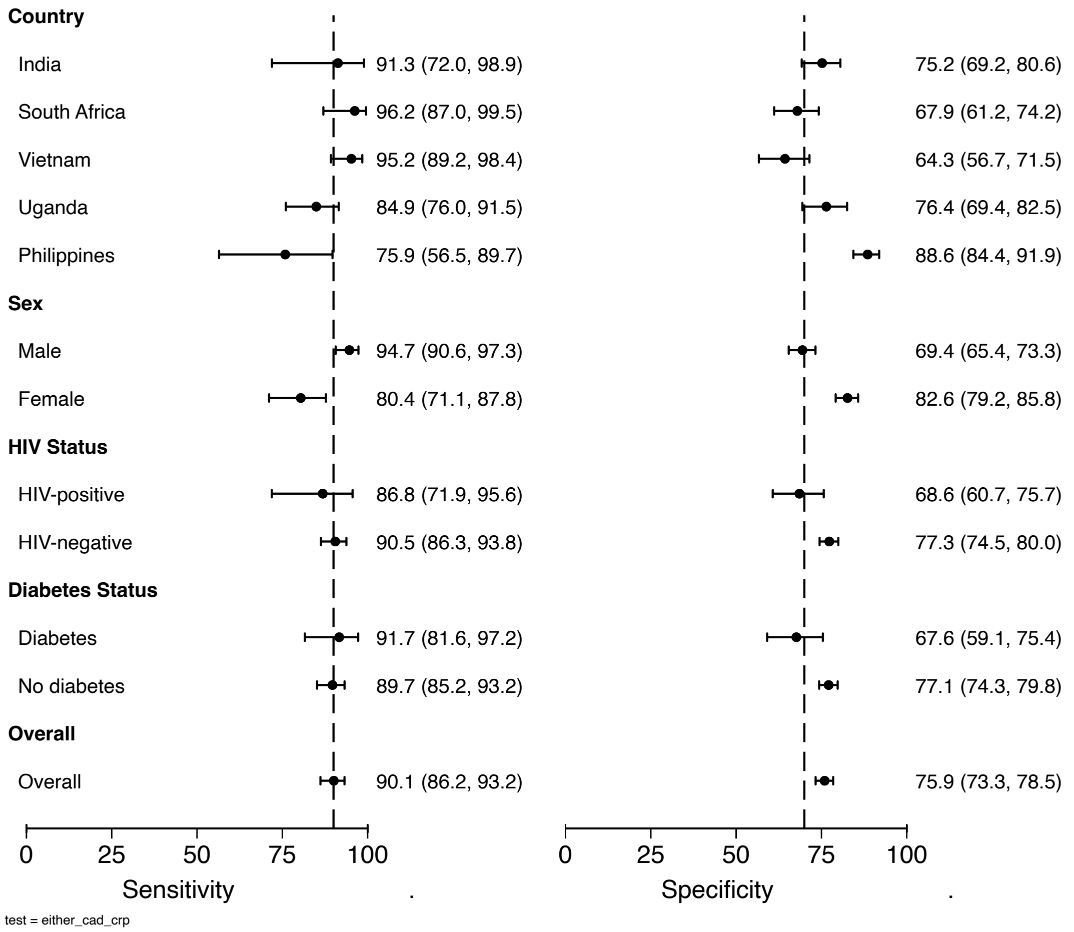

**Figure S6. Subgroup analysis of two-step screening algorithm CRP-CAD4TB using a sequential negative serial screening algorithm.**

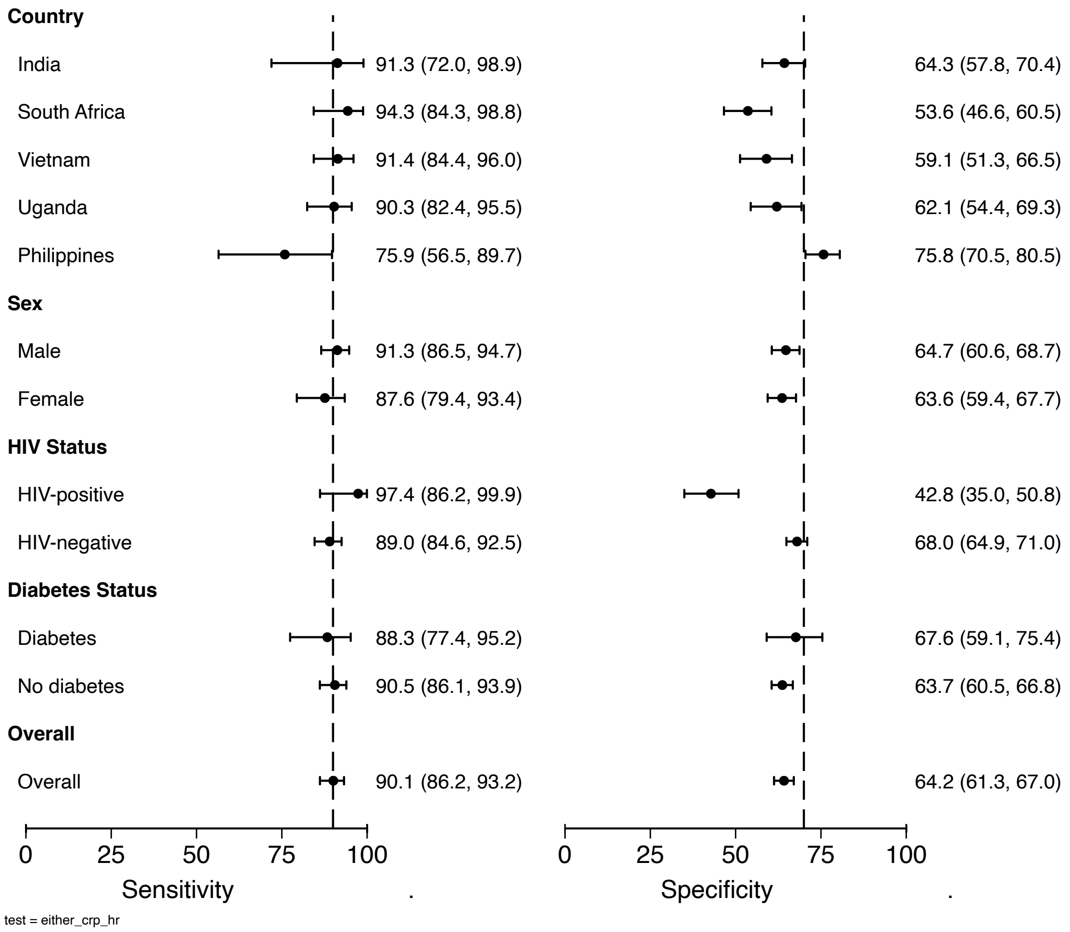

**Figure S7. Subgroup analysis of two-step screening algorithm CRP-Xpert HR using a sequential negative serial screening algorithm.**

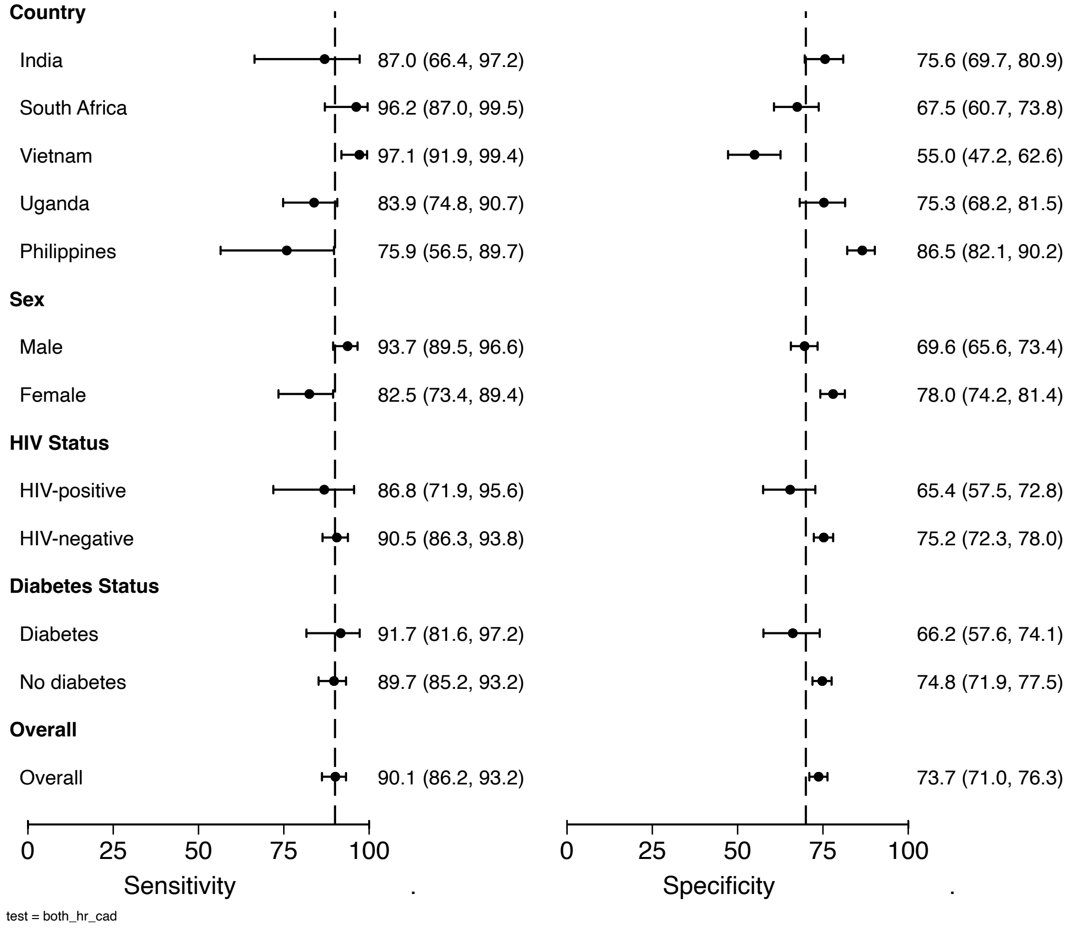

**Figure S8. Subgroup analysis of two-step screening algorithm Xpert HR-CAD using a sequential negative serial screening algorithm.**

**Table S7. Agreement between binary triage tests (percent agreement, Cohen's Kappa statistic)**

| Percent agreement (Kappa) | CAD4TB | Xpert HR | CRP | CAD-Xpert HR (sequential negative) | CAD-CRP (sequential negative) | CRP-Xpert HR (sequential negative) | CAD-Xpert HR (sequential positive) | CAD-CRP (sequential positive) | CRP-Xpert HR (sequential positive) |
|---|---|---|---|---|---|---|---|---|---|
| **CAD4TB** | - | 69.0% (0.38) | 65.5% (0.33) | 83.3% (0.65) | 92.0% (0.84) | 70.0% (0.40) | 92.0% (0.84) | 99.4% (0.99) | 69.0% (0.37) |
| **Xpert HR** | - | - | 69.4% (0.39) | 76.2% (0.51) | 71.8% (0.43) | 97.4% (0.95) | 75.1% (0.50) | 69.4% (0.38) | 99.4% (0.99) |
| **CRP** | - | - | - | 66.1% (0.36) | 66.8% (0.36) | 71.1% (0.43) | 67.0% (0.36) | 65.8% (0.33) | 69.8% (0.40) |
| **CAD-Xpert HR (sequential negative)** | - | - | - | - | 88.5% (0.75) | 77.0% (0.53) | 83.5% (0.65) | 82.9% (0.64) | 75.7% (0.51) |
| **CAD-CRP (sequential negative)** | - | - | - | - | - | 73.9% (0.47) | 88.9% (0.77) | 91.8% (0.83) | 71.6% (0.43) |
| **CRP-Xpert HR (sequential negative)** | - | - | - | - | - | - | 75.7% (0.51) | 70.6% (0.41) | 97.1% (0.94) |
| **CAD-Xpert HR (sequential positive)** | - | - | - | - | - | - | - | 92.5% (0.85) | 75.1% (0.50) |
| **CAD-CRP (sequential positive)** | - | - | - | - | - | - | - | - | 69.3% (0.38) |
| **CRP-Xpert HR** | - | - | - | - | - | - | - | - | - |

| |
|---|
| **(sequential positive)** |
